# Efficacy and Safety of Transcranial Temporal Interference Stimulation for Treating Bipolar Disorder with Depressive Episodes

**DOI:** 10.1101/2024.11.19.24317540

**Authors:** Hetong Zhou, Minmin Wang, Shuangyu Qi, Qianfeng Chen, Jianbo Lai, Zhengping Wu, Ruobing Liu, Liang Wang, Junxian Tang, Shaomin Zhang, Shaohua Hu

## Abstract

**Background:** Bipolar disorder (BD) presents significant challenges in clinical management, often characterized by persistent depressive symptoms and cognitive deficits. Transcranial temporal interference stimulation (tTIS) has shown promise in targeting deep brain structures with minimal invasiveness. This study aimed to assess the efficacy and safety of tTIS in newly diagnosed or medication-washout BD patients.

**Methods:** We conducted a single-arm clinical trial with 36 BD patients who underwent 10 sessions of tTIS targeting the left nucleus accumbens over one week, with two sessions per day. Each tTIS session lasted 20 minutes, utilizing a maximum current intensity of 2 mA and an envelope stimulation frequency of 40 Hz. Depressive symptoms were assessed pre- and post-intervention using validated scales, including HAMD, QIDS, MADRS, and HAMA, while cognitive functions were evaluated using standardized neuropsychological tests. Safety was monitored through adverse event reporting.

**Results:** Among 36 participants, 25 completed the full intervention protocol. Significant reductions were observed in depressive symptom scores: HAMD decreased from 23.36 to 16.16 (*P* < 0.0001), QIDS from 13.52 to 9.68 (*P* < 0.001), MADRS from 39.12 to 31.28 (*P* < 0.01), and HAMA from 19.68 to 15.44 (*P* < 0.05). Cognitive evaluations demonstrated notable improvements in memory and executive function. Adverse events were mild, primarily limited to transient scalp discomfort.

**Conclusions:** This study provides preliminary evidence supporting the efficacy and safety of tTIS in alleviating depressive symptoms and improving cognitive function in BD patients.

## 1. Introduction

Bipolar disorder (BD) is a psychiatric condition characterized by recurrent episodes of depression, mania, or hypomania^1^. Among most individuals with BD, depressive symptoms are more predominant than manic symptoms. Bipolar depression has been shown to account for a significant proportion of the overall morbidity and mortality associated with BD. Notably, BD is linked to a markedly elevated risk of suicide, with the risk during depressive episodes estimated to be 20–30 times higher than in the general population^2^. Current pharmacological interventions for bipolar depression have limited efficacy improvements and are often associated with side effects^3^. Hence, there is an urgent need to explore noninvasive, safe, and rapid-acting therapeutic approaches for treating bipolar depression.

Traditional noninvasive brain stimulation technique for BD, such as transcranial direct current stimulation (tDCS)^4^, transcranial alternating current stimulation (tACS)^5,6^, and transcranial magnetic stimulation (TMS)^7^, have demonstrated promising clinical outcomes. However, these techniques primarily target cortical regions, while emerging evidence suggests that abnormalities in brain functional networks in BD patients involve several subcortical structures. For instance, the severity of anhedonia in BD has been strongly linked to dysfunction in the striatum and prefrontal cortex^8,9^. Previous studies have revealed abnormal activation of the nucleus accumbens (NAcc) in BD, which is a critical region involved in reward processing^10,11^. Research has further explored the relationship between the NAcc and the ventromedial prefrontal cortex (vmPFC) in BD patients, showing decreased functional connectivity between the striatum and prefrontal cortex during reward anticipation compared to healthy controls^12^. Additionally, resting-state functional connectivity between the NAcc and vmPFC is increased in BD patients^13,14^. Pharmacological studies have also indicated that the NAcc exhibits abnormal local morphology in BD patients, which can be modulated by medication to stabilize mood^15,16^. Therefore, noninvasive interventions targeting subcortical structures like the NAcc may offer superior therapeutic benefits for BD patients.

Transcranial temporal interference stimulation (tTIS) is an emerging noninvasive brain stimulation technique that has been developed to modulate neuronal activity in deep brain regions^17^. The fundamental principle of tTIS involves the application of two high-frequency transcranial alternating current stimulation waveforms with slightly offset frequencies^18,19^. When these high-frequency signals are applied to a target brain region, their interaction generates a low-frequency amplitude-modulated waveform, or “envelope,” within the stimulated tissue^20,21^. tTIS has been preliminarily explored in clinical applications such as Parkinson’s disease and Alzheimer’s disease, with basic research confirming its potential to modulate activity in deep brain regions^22-25^. However, the clinical efficacy and safety of tTIS in BD patients remain underexplored.

This study aims to investigate the efficacy and safety of tTIS in treating depressive episodes in first-diagnosed bipolar disorder patients. By targeting the NAcc using tTIS, this research seeks to evaluate its potential as a noninvasive, rapid-acting, and effective therapeutic approach for bipolar disorder. The findings of this study could provide valuable insights into novel treatment strategies for BD and contribute to advancing noninvasive neuromodulation techniques for psychiatric disorders.

## 2. Methods and Materials

### 2.1 Trial Design

In this study, a single-arm clinical trial designed to investigate the efficacy and safety of tTIS in treating depressive episodes in patients with bipolar disorder. The participants were recruited from the First Affiliated Hospital of Zhejiang University School of Medicine. The trial was approved by the Institutional Review Board of First Affiliated Hospital of Zhejiang University School of Medicine and conducted in compliance with the ethical principles of the Declaration of Helsinki. The trial was registered at ClinicalTrials.gov (NCT06516991). Written informed consent was obtained from all participants prior to enrollment.

After enrollment, participants underwent standardized tTIS intervention targeting the left nucleus accumbens. Each treatment session lasted 20 minutes, with a maximum output current of 2 mA adjusted based on individual head models. The two pairs of electrical stimulation frequencies in tTIS are 1000 Hz and 1040 Hz, respectively. Sessions were conducted twice daily, Monday through Friday, for a total of 10 sessions. Before each session, participants were shown a 5-minute emotion-arousing video to standardize their baseline emotional state. No medication intervention is used during the intervention process.

### 2.2 Participants

We recruited patients aged 16-50 years who met the Diagnostic and Statistical Manual of Mental Disorders, Fifth Edition (DSM-5) diagnostic criteria for bipolar disorder. The inclusion criteria were as follows: 17-item Hamilton Rating Scale for Depression (HRDS-17) >17 ; Subjects had never received psychiatric medication or had undergone a washout period prior to enrollment; The subject/legal guardian is willing to actively cooperate with the treatment and sign an informed consent form after being fully informed of tTIS. Patients who met the following criteria were excluded: Co-morbid other psychiatric disorders, including anxiety spectrum disorders, mental retardation, substance dependence, etc.; history of a serious physical illness or a disease that may affect the central nervous system; neurologic disorders or risk of seizures such as previous cranial disorders, head trauma, abnormal electroencephalograms, magnetic resonance evidence of structural brain abnormalities, or a family history of epilepsy; contraindications to magnetic resonance scanning or tTIS, such as the presence of metallic or electronic devices in the body (intracranial metallic foreign bodies, cochlear implants, pacemakers and stents, and other metallic foreign bodies); have received or are receiving electroconvulsive therapy, modified electroconvulsive therapy, transcranial magnetic stimulation, transcranial direct current stimulation, transcranial alternating current stimulation, or other neurostimulation treatments; pregnant and lactating women, and women of childbearing age with positive urine pregnancy.

### Treatment Protocol of Transcranial Temporal Interference Stimulation

To achieve envelope-specific stimulation of the left NAcc, customized electrode configurations were developed for each participant using individual magnetic resonance imaging (MRI) data. The implementation involved the following steps:

#### Step 1: Individual Head Model Construction

Individual head models were constructed to simulate the electric field distribution under various stimulation configurations. The forward model was developed using the CHARM tool, which processes T1-weighted MRI scans to perform tissue segmentation, cortical reconstruction, and meshing. The head model included 10 different tissue types: white matter, gray matter, cerebrospinal fluid, bone, scalp, eyes, blood, compact bone, spongy bone, and muscle. Each tissue was assigned a specific conductivity value to ensure accurate modeling.

#### Step 2: *Leadfield* Calculation

The *leadfield* matrix was calculated to estimate the electric field distribution generated by a given current input through specific electrode configurations. The initial calculation was performed in a surface-*based* source space, decomposing the brain tissues into thousands of tetrahedra. The leadfield was then resampled into a volume-based source space, comprising 1 mm^3^voxels.

For this study, 66 electrode positions from the 10-10 EEG system were used for *leadfield* computation, with Cz as the reference electrode. To enhance participant comfort during MRI scanning, electrode positions on the occipital region were excluded.

#### Step 3: Optimal Electrode Montage Selection

The efficacy of tTIS depends on maximizing the envelope electric field 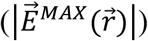 in the target region while suppressing its effects in non-target areas. The envelope electric field 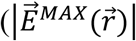 was computed using the following formula:

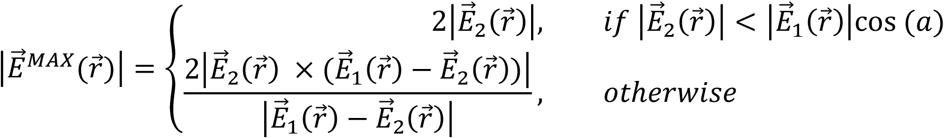

Where 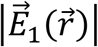and 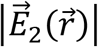represent the maximum amplitudes of the two electric fields generated by two electrode pairs: *C*_11_ − *C*_12_ (current *I*_1_) and *C*_21_ − *C*_22_ (current *I*_2_). The respective fields were calculated using the following equations:

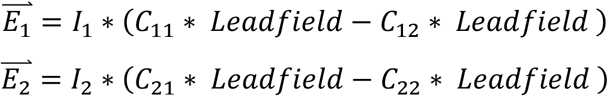

Given the 66 electrode positions, there were 4,324,320 potential configurations. Full-brain voxel calculations were computationally intensive, so this study adopted a targeted approach to optimize efficiency.

#### Target Region Screening

All possible configurations were evaluated within the target region, significantly reducing computational time. A 0.4 V/m threshold was applied to select electrode combinations based on the maximum envelope electric field in the target region. Among the remaining combinations, the top 10% with the highest average field intensity were selected.

#### Non-Target Region Suppression

The envelope electric field in gray matter regions was calculated for the shortlisted combinations. Configurations with minimal non-target field intensity was prioritized to enhance stimulation focus on the target area.

Once the optimal tTIS parameters (electrode positions and stimulation currents) were determined, the stimulation was administered using a tTIS device (SHTIS, Shanhai Medical Ltd, Nanjing, China) and an EEG cap to position electrodes accurately Ag/AgCl electrodes with hydrogel were used to ensure good conductivity and minimize skin impedance.

### 2.4 MRI Scanning

The MRI data were collected using a 3.0 Tesla scanner (GE, SIGNA) at the First Affiliated Hospital, Zhejiang University School of Medicine. High-resolution T1-weighted images were acquired using a sagittal three-dimensional turbo-flash imaging sequence with the following parameters: TR = 2300 msec, TE = 2.32 msec, FA = 8 degrees, FOV = 240 × 240 mm ^2^, and slice thickness = 0.90 mm. Resting-state functional MRI images were obtained using a single-shot gradient-recalled echo-planar imaging sequence in the axial plane parallel to the anterior-posterior commissure line, with parameters including 52 slices, TR = 1000 msec, TE = 34 msec, FA = 50 degrees, FOV = 230 × 230 mm ^2^, and slice thickness = 2.5 mm.

### 2.5 Clinical Assessment

The primary outcome was the change in the patient’s HAMD score before and after treatment. Secondary endpoints included: changes in patients’ scores on the 14 item Hamilton Anxiety Scale (HAMA-14), Montgomery-Asberg Depression Rating Scale (MADRS), and Young Mania Rating Scale (YMRS) before and after treatment; the proportions of responders (defined with a reduction of 50 % or more from baseline in the HAMD-17 total score) after intervention; changes in patients’ pleasure deficit symptoms before and after treatment assessed using the Snaith-Hamilton Pleasure Scale (SHAPS) and Temporal Experience of Pleasure Scale (TEPS); changes in Self-Rating Anxiety Scale (SAS) and 16-item Quick Inventory of Depressive Symptomatology Self-Report (QIDS-SR16) before and after treatment. In addition, we used the THINC-integrated tool (THINC-it) to assess changes in cognitive functioning before and after treatment in patients.

The collected scores from the assessment tools were subjected to statistical analysis using paired t-tests. Paired t-tests were used to investigate specific changes between baseline and post-treatment assessments.

## 3. Results

### 3.1 Clinicodemographic Characteristics of Patient

A total of 36 participants were screened for this study, of whom 9 were excluded due to eligibility criteria. The final sample included 27 patients diagnosed with bipolar disorder. Among these, 25 participants completed the full protocol of 10 planned tTIS intervention sessions and underwent pre- and post-intervention assessments. The primary reasons for incomplete trial participation included shorter hospitalization periods, scheduling conflicts, and clinical improvement leading to discharge.

### 3.2 Emotion Symptoms

Emotion symptoms were assessed using validated rating scales before and after the tTIS intervention. As shown in Figure 1, among the 25 participants who completed the full intervention protocol, significant improvements were observed in overall scores on the HAMD, QIDS, MADRS, and HAMA (all *P* < 0.05). Specifically: The mean HAMD score decreased from 23.36 (pre-intervention) to 16.16 (post-intervention). The QIDS score showed a marked reduction from 13.52 (pre-intervention) to 9.68 (post-intervention). The MADRS score improved significantly, decreasing from 39.12 (pre-intervention) to 31.28 (post-intervention). The HAMA score also demonstrated a significant reduction, dropping from 19.68 (pre-intervention) to 15.44 (post-intervention). Although the Young Mania Rating Scale (YMRS) did not exhibit statistically significant changes, there was a reduction in the mean score from 10.4 (pre-intervention) to 7.72 (post-intervention). These results suggest that tTIS intervention effectively alleviates depressive and anxiety symptoms in participants, while also modestly impacting manic symptoms.

**Figure 1.**
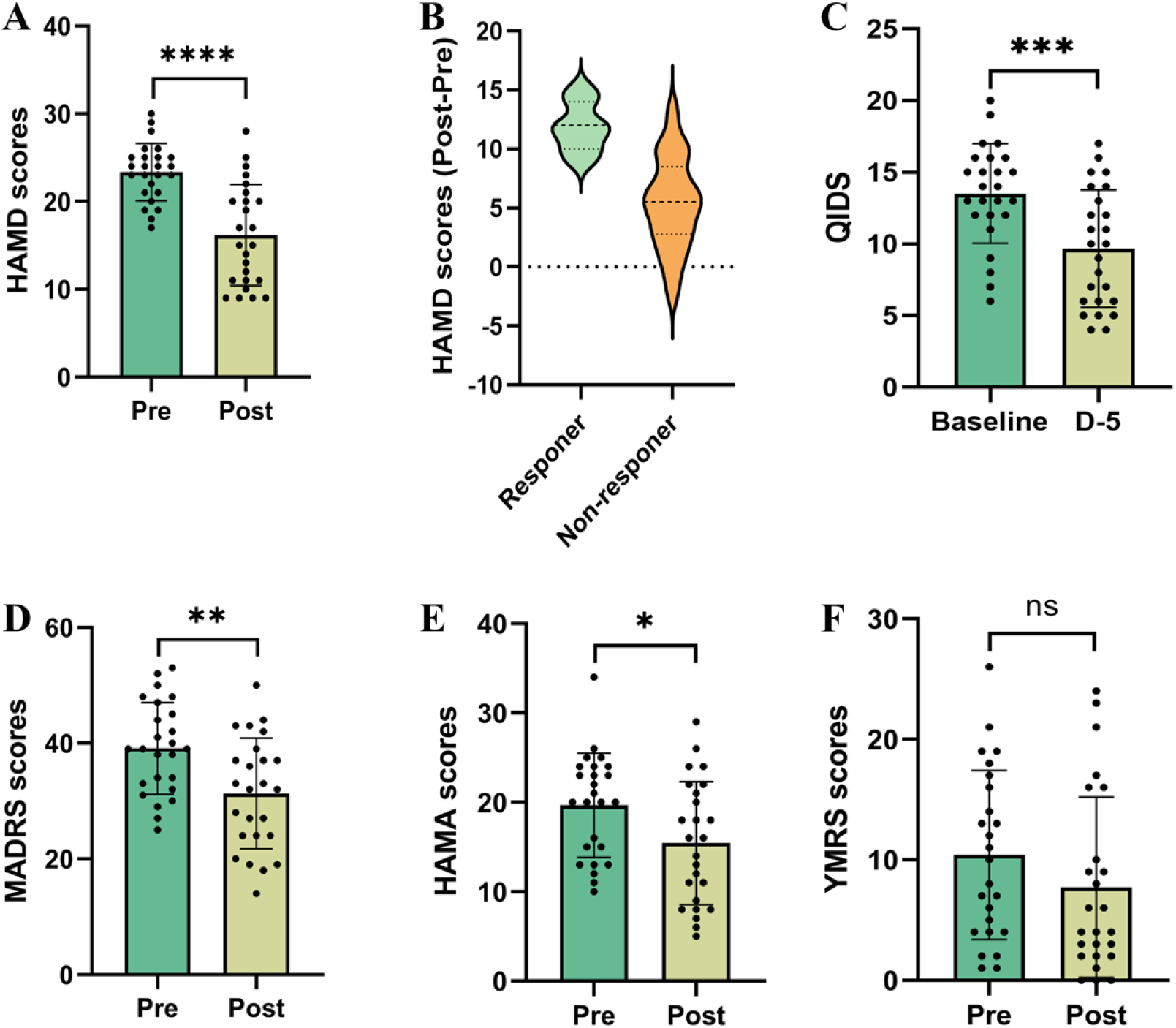
Effects of tTIS Intervention on Depressive and Anxiety Symptoms. (A) Results of HAMD scores before and after tTIS intervention. (B) Reduction rates in HAMD scores, comparing responders (defined as participants with a reduction of 50% or more from baseline in the HAMD-17 total score) and non-responders. (C) QIDS scores before and after tTIS intervention. (D) MADRS scores before and after tTIS intervention. (E) HAMA scores before and after tTIS intervention. (F) YMRS scores before and after tTIS intervention. Statistical significance is indicated as follows: \**P* < 0.05, \*\**P* < 0.01, \*\*\**P* < 0.0001.

### 3.3 Cognitive Function

Cognitive function was evaluated using the THINC-it tool before and after the tTIS intervention. Figure 2 illustrates the changes in cognitive performance scores before and after the intervention. Post-intervention scores demonstrated a trend toward improvement in PDQ-5, CRT, DSST, and N-back scores. Among these, statistically significant differences were observed in DSST and N-back reaction time (NbackRT) scores (*P* < 0.05), highlighting measurable cognitive enhancement following the intervention. These improvements suggest that tTIS intervention may provide potential neurocognitive benefits.

**Figure 2.**
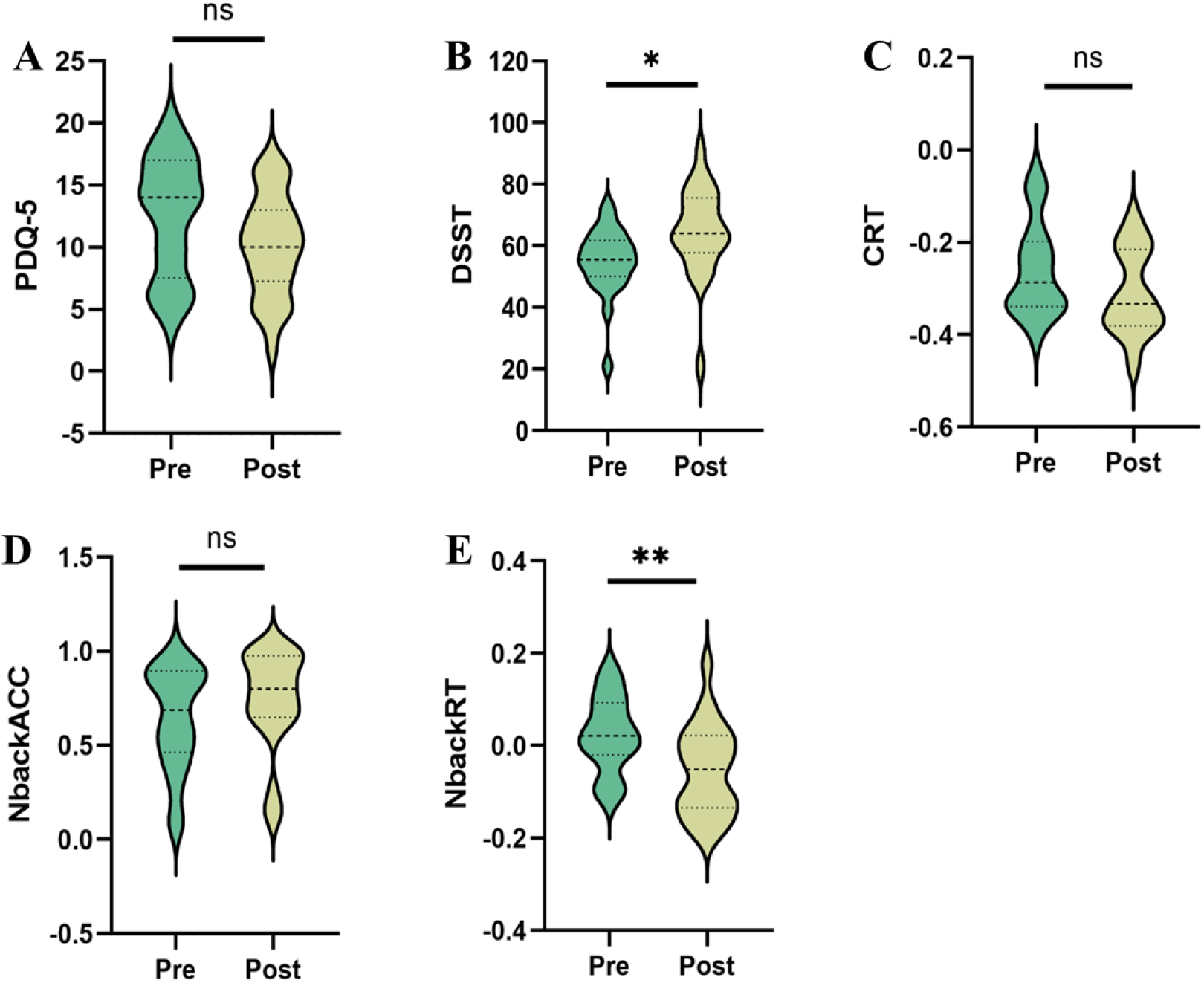
Cognitive Improvement Following tTIS Intervention.(A) Pre- and post-intervention PDQ-5 scale scores; (B) Pre- and post-intervention DSST scores; (C) Pre- and post-intervention CRT scores; (D-E) Pre- and post-intervention N-back task accuracy (N-backACC) and reaction time (N-backRT) scores. Statistical significance is indicated as follows: \**P* < 0.05, \*\**P* < 0.01.

### 3.4 Adverse Effects

Adverse events were minor and transient. During the initial ramp-up phase of the intervention, participants commonly reported mild local discomfort at the stimulation sites, which resolved quickly without requiring intervention. Additionally, one participant experienced a mild headache attributed to a cold, which subsided with symptomatic treatment. No severe or lasting adverse events were reported throughout the study, indicating the safety and tolerability of the tTIS intervention.

## 4. Discussion

This study represents a significant step forward in the exploration of tTIS as a therapeutic intervention for bipolar disorder. For the first time, we have demonstrated the efficacy and safety of tTIS in improving depressive symptoms and cognitive function in newly diagnosed or drug-washed BD patients. These findings hold substantial promise for advancing neuromodulation techniques in psychiatric disorders.

Our results showed a notable reduction in depressive symptoms following tTIS intervention targeting the NAcc. This is consistent with the role of the NAcc in the reward system and its dysfunction in depression. Previous studies have indicated that electrical stimulation of the NAcc can modulate neural circuits implicated in mood regulation, potentially alleviating depressive symptoms ^26,27^. The observed improvements in our study extend this understanding by employing a noninvasive approach with tTIS, which leverages spatially focused stimulation to achieve therapeutic effects. These findings underscore the potential of tTIS as an alternative to invasive neuromodulation methods, such as deep brain stimulation, with broader accessibility and fewer risks.

In addition to mood improvement, our study demonstrated significant enhancements in cognitive functions, including attention, memory, and executive function, following the tTIS intervention. This aligns with emerging evidence linking neuromodulation of the NAcc to cognitive improvements ^28,29^. The mechanism underlying this effect may involve increased functional connectivity between the NAcc and prefrontal cortical regions, which are critical for cognitive processes. Future studies employing neuroimaging could further elucidate these pathways and validate the observed cognitive benefits.

While our study provides valuable insights into the potential benefits of tTIS for improving neuropsychiatric outcomes in BD, several limitations must be considered. Firstly, this was a single-arm study focusing on drug-naïve or drug -washed BD patients, with no concurrent pharmacological treatment during the intervention period. Although this design minimized confounding factors, a randomized controlled trial (RCT) with a sham stimulation arm and larger patient cohorts is necessary to establish causality and generalizability. Multicenter RCTs could further enhance the robustness of our findings. Secondly, our sample size, while sufficient to demonstrate statistical significance, was moderate. Including a larger and more diverse patient population in future studies would improve the generalizability of our results and allow for subgroup analyses based on demographic or clinical characteristics. Last, although tTIS showed immediate post-intervention efficacy, we were unable to assess its long-term effects due to the transition to pharmacological treatments after the intervention. Future research incorporating extended follow-up periods could provide critical insights into the durability of tTIS-induced benefits and its role in long-term BD management.

## 5. Conclusion

In summary, this study demonstrate that tTIS is a feasible, safe, and potentially effective intervention for depressive symptoms in BD patients, with additional benefits in cognitive function and minimal adverse effects.

## Data Availability

Data involved in this study are available upon reasonable request.

## Data availability

Data involved in this study are available upon reasonable request.

### Acknowledgments

We would like to extend our heartfelt gratitude to all the patients who participated in this clinical study and their families for their invaluable support.

